# Decline in COPD Admissions During the COVID-19 Pandemic Associated with Lower Burden of Community Respiratory Viral Infections

**DOI:** 10.1101/2021.05.18.21257383

**Authors:** Jennifer Y So, Nathan N O’Hara, Blaine Kenaa, John G Williams, Christopher L deBorja, Julia F Slejko, Zafar Zafari, Michael Sokolow, Paul Zimand, Meagan Deming, Jason Marx, Andrew N Pollak, Robert M Reed

## Abstract

**Background:** The COVID-19 pandemic has led to widespread implementation of public health measures, such as stay-at-home orders, social distancing, and masking mandates. In addition to decreasing spread of SARS-CoV2, these measures also impact the transmission of seasonal viral pathogens, which are common triggers of COPD exacerbations. Whether reduced viral prevalence mediates reduction in COPD exacerbation rates is unknown.

**Methods:** We performed retrospective analysis of data from a large, multicenter healthcare system to assess admission trends associated with community viral prevalence and with initiation of COVID-19 pandemic control measures. We applied difference-in-differences (DiD) analysis to compare season-matched weekly frequency of hospital admissions for COPD before and after implementation of public health measures for COVID-19. Community viral prevalence was estimated using regional Center for Disease Control and Prevention test positivity data and correlated to COPD admissions.

**Results:** Data involving 4,422 COPD admissions demonstrated a season-matched 53% decline in COPD admissions during COVID-19 pandemic, which correlated to community viral burden (r=0.73; 95% CI: 0.67 to 0.78) and represented a 36% greater decline over admission frequencies observed in other medical conditions less affected by respiratory viral infections (IRR, 0.64; 95% CI, 0.57 to 0.71, p<0.001). The post-COVID-19 decline in COPD admissions was most pronounced in patients with fewer comorbidities and without recurrent admissions.

**Conclusion:** The implementation of public health measures during the COVID-19 pandemic was associated with decreased COPD admissions. These changes are plausibly explained by reduced prevalence of seasonal respiratory viruses.

## Introduction

Prior to the COVID-19 pandemic, chronic obstructive pulmonary disease (COPD) was the fourth-leading cause of death worldwide and a common cause of hospital admissions in the United States.^1-4^ COPD exacerbations cause a staggering burden of morbidity, mortality, and cost to the health care system.^5-7^

The recent COVID-19 pandemic has led to a significant interruption in health care delivery, with reduced admissions for COPD and other non-COVID illnesses.^8-13^ This is due to a variety of factors, including avoidance of crowded hospitals due to fear of contracting SARS-CoV2 (the virus responsible for COVID-19), reallocation of hospital resources to treat rising number of COVID-19 patients, and utilization of telemedicine to manage chronic diseases at home. There is compelling evidence indicating that public health measures (such as stay at home orders, social distancing, and masking mandates with strict limitations on large gatherings to curb the spread of COVID-19) are also effective at decreasing the transmission of seasonal viral pathogens.^14,15^

It is plausible that measures taken to reduce the spread of SARS-CoV2 have also resulted in reductions in COPD exacerbations, due in large part to reduced burden of non-SARS-CoV2 community virus transmission. Respiratory viral infections are the most commonly identified trigger for COPD exacerbations. They are implicated in approximately 50% of cases, and result in a disease course that tends to be more severe and longer in duration.^16,17, 18^

We aimed to determine whether public health measures related to the COVID-19 pandemic were associated with reduced seasonal viral infections and COPD-related hospital admissions. Using data from a large, multicenter healthcare system, we hypothesized that the public health measures related to the COVID-19 pandemic decreased hospital admissions for COPD exacerbations.

## Materials and Methods

### Study Design and Data Sources

We conducted a difference-in-differences analysis of retrospective data to compare weekly hospital admissions for COPD pre-versus post-COVID-19 public health measures. Stay-at-home mandate and masking mandate were instituted in the state of Maryland on March 30^th^,2020 and April 18^th^, 2020, respectively. April 1^st^, 2020 was selected as the start of post-COVID-19 period. A difference-in-differences model estimates the average effect among an exposure group while controlling for factors unrelated to the exposure of interest that would affect both exposed and unexposed groups. In this case, a key difference between the groups is the association to respiratory viral infections as triggers for destabilization and subsequent hospitalization. We obtained hospital data from the University of Maryland Medical System, which consists of 12 hospitals throughout the state of Maryland. Patients with a primary diagnosis of COPD for their hospital admission were identified using *International Classification of Diseases, Tenth Revision, Clinical Modification* codes (ICD-10-CM) (Online supplement, Table 1a). We selected a control group of primary diagnoses that would account for a similar reservation among patients to seek hospital care due to a fear of COVID-19 transmission. The control group included all admissions with a primary diagnosis of myocardial infarction, diabetes mellitus, or congestive heart failure, and were identified using ICD-10-CM codes (Online supplement, Table 1b). The control group diagnoses were also selected for informative epidemiologic aspects. Both congestive heart failure and myocardial infarction admission rates experience seasonal variability and are associated with trends in viral infections, although less robustly than observed in COPD.^19-22^ As such, selection of these diagnoses was an intentionally conservative approach. Diabetes, on the other hand, demonstrates little seasonality, although it warrants emphasis that all analyses were seasonally controlled. Only the patients with the above ICD-10-CM codes were included in the study. The analyses were restricted to only include data from April 1 through September 30 in 2018 through 2020 to ensure a seasonal match. The period prior to implementation of public health measures included data from 2018 and 2019, and the period after implementation included data from 2020. The study was approved by the University of Maryland institutional review board (HP-00093452).

### Outcomes

The primary outcome was weekly hospital admissions. Secondary outcomes included weekly counts of in-hospital mortality, severe exacerbations (identified as requiring intensive care unit admission), and hospital length of stay. Additionally, we calculated community viral percent positivity to assess the correlation between viral trends and COPD admissions. Data on respiratory virus incidence was provided by the Centers for Disease Control and Prevention’s (CDC) National Respiratory and Enteric Virus Surveillance System and the CDC FluView report. Respiratory virus results are reported weekly from participating laboratories, and the number of tests and positive tests for the included respiratory viruses were provided from the middle Atlantic and South Atlantic census regions encompassing January 1, 2018 through October 1, 2020. Viruses reported included RSV, human metapneumovirus, seasonal coronaviruses (including 229E, HKU1, NL63, and OC43), human parainfluenzavirus (types 1-3), rhinovirus, and influenza.^23^

### Ascertainment of Patient Characteristics

Patient characteristics were extracted from the electronic health records to assess for any imbalance between time periods or exposure groups. We also used ICD-10-CM codes to generate Charlson Comorbidity Index scores, and diagnoses of morbid obesity, tobacco use, alcohol-related disorders, substance-related disorders, and social challenges (Online supplement, Table 1c-d).

### Statistical Analysis

The primary analysis of the change in weekly hospital admissions applied a negative binomial model, adjusting for an observed imbalance between periods in patient age and a weekly time trend. We included a dummy variable for the admitting hospital as a random effect in the model to account for variation in public health measure adherence and socioeconomic compositions among the catchment areas. We used Pearson’s correlation coefficients to evaluate the association between community viral percent positivity data with admissions for COPD, congestive heart failure, diabetes mellitus, and myocardial infarction.

The in-hospital mortality and severe exacerbation models were fit using negative binomial regression. We used a quantile regression model to evaluate the effect of the COVID-19-related public health measures on the median hospital length of stay days among COPD patients. In all models, we adjusted for patient age and accounted for the hospital as a random effect.

We performed two subgroup analyses to assess heterogeneity of effect based on co-morbidities and recurrent admissions. The comorbidities were identified at the time of admission, and reported using the Charlson Comorbidity Index (CCI) which was stratified between patients with a CCI ≤3 and patients with a CCI >3 (Online supplement 1c).^24^ Patients were classified as having recurrent admissions if they had more than one admission for the same primary diagnosis during the study period. The subgroup term was interacted with the comparison groups and exposure for a difference-in-difference-in-differences model. With this approach, we accounted for the subgroup covariate as an interaction term to the difference-in-differences equation. This model ascertained if the difference-in-differences estimate changed between subgroups.

We performed two sensitivity analyses. In the first analysis, we added patients who had a primary admission diagnosis of COVID-19 and a secondary diagnosis of COPD (Online supplement 1e) to the COPD group. The intention of this analyses was to determine if the decline in COPD admissions was due to related COVID-19 diagnosis taking precedence in the diagnosis. In the second analysis, we rebuilt three versions of our primary difference-in-differences model, one model for each control group’s primary admission diagnosis - congestive heart failure, diabetes mellitus, or myocardial infarction. The purpose of this sensitivity analyses was to evaluate if our estimates were robust across the various control groups.

Difference-in-differences estimates for the negative binomial models are reported as an incidence rate ratio with 95% confidence intervals. The difference-in-differences estimates for the quantile regression model is reported as the change in median days of hospital stay with 95% confidence intervals. Confidence intervals were calculated using bootstrap methods.

The analyses were performed with R Version 4.0.0 (Vienna, Austria). For the primary analysis, we considered a two-tailed p < 0.05 as statistically significant. We did not adjust for multiple testing, and all secondary outcomes should be considered exploratory. We did not impute for missing data.

## Results

There were 18,587 admissions during the specified periods; 13,695 occurred prior to the implementation of COVID-19-related public health measures (pre-COVID-19), and 4,892 observations occurred afterwards (post-COVID-19). There were 4,422 hospital admissions for COPD, 6,414 for congestive heart failure, 4,217 for diabetes, and 3,534 for myocardial infarction.

The characteristics for the COPD patients and the control group during the periods under consideration are provided in Table 1. Notably, the average age of COPD patients admitted during the post-COVID-19 period was older (62.7 ± 16.5 years) compared to pre-COVID-19 period (55.3 ± 24.4 years). There were no significant differences in gender, race, or distribution of Charlson Comorbidity Index scores. The mean age of all patients with COPD admission was 57 ± 23 years, and more than half were female (n=2,499, 57%). The majority of these patients had three or fewer comorbidities (n=3683, 83%).

### Primary Outcome

The post—COVID crude weekly COPD admissions decreased by 53% compared to the pre-COVID-19 seasonally-comparable period (31.5 vs. 67.4 admissions/week respectively). Compared to declines observed in other medical conditions, weekly COPD admissions demonstrated a 36% greater decline (incidence rate ratio (IRR), 0.64; 95% CI, 0.57 to 0.71, p<0.001) (Table 2, Figure 1). To ensure that the results were not attributable to concomitant SARS-CoV2 infection, a sensitivity analysis was performed including admissions for primary diagnosis of COVID-19 with a secondary diagnosis of COPD included in the COPD group. The findings remained significant even with this conservative approach (Online supplement, Table 3). The results were also consistent within subsets of the control group admitted for congestive heart failure (IRR, 0.66; 95% CI, 0.59 to 0.75), diabetes mellitus (IRR, 0.57; 95% CI, 0.51 to 0.65), and myocardial infarction (IRR, 0.69; 95% CI, 0.60 to 0.77) (Online supplement Table 4). In addition, there was an apparent temporal correlation between weekly COPD admissions and community viral percent positivity (r, 0.73; 95% CI: 0.67 to 0.78) (Figure 2).

**Figure 1.**
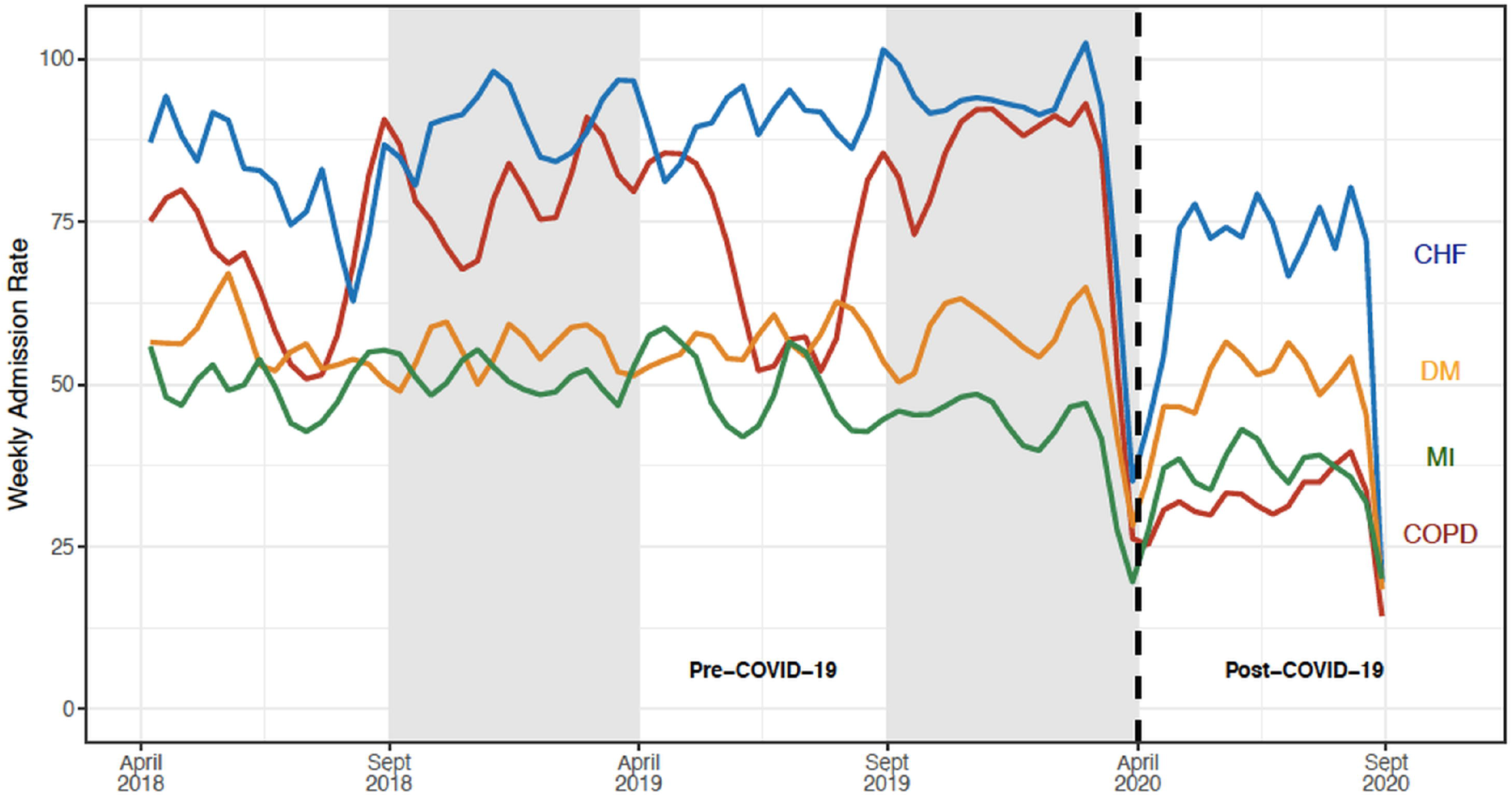
Weekly Hospital Admission Rates for Chronic Obstructive Pulmonary Disease, Congestive Heart Failure, Diabetes Mellitus, and Myocardial Infarction. Our analyses compared the post-intervention period (April 1, 2020 through September 30, 2020) with a season-matched pre-intervention period comprising of April 1, 2018 through September 30, 2018 and April 1, 2019 through September 30, 2019. The grey shaded areas denote data that were not included in the season-matched analysis. The pandemic time period was associated with a 53% decline in COPD admissions (from 67.4 to 31.5 admissions/week) which represented a 36% greater decline than that observed in other medical indications evaluated (IRR, 0.64; 95% CI, 0.57 to 0.71, p<0.001).

**Figure 2.**
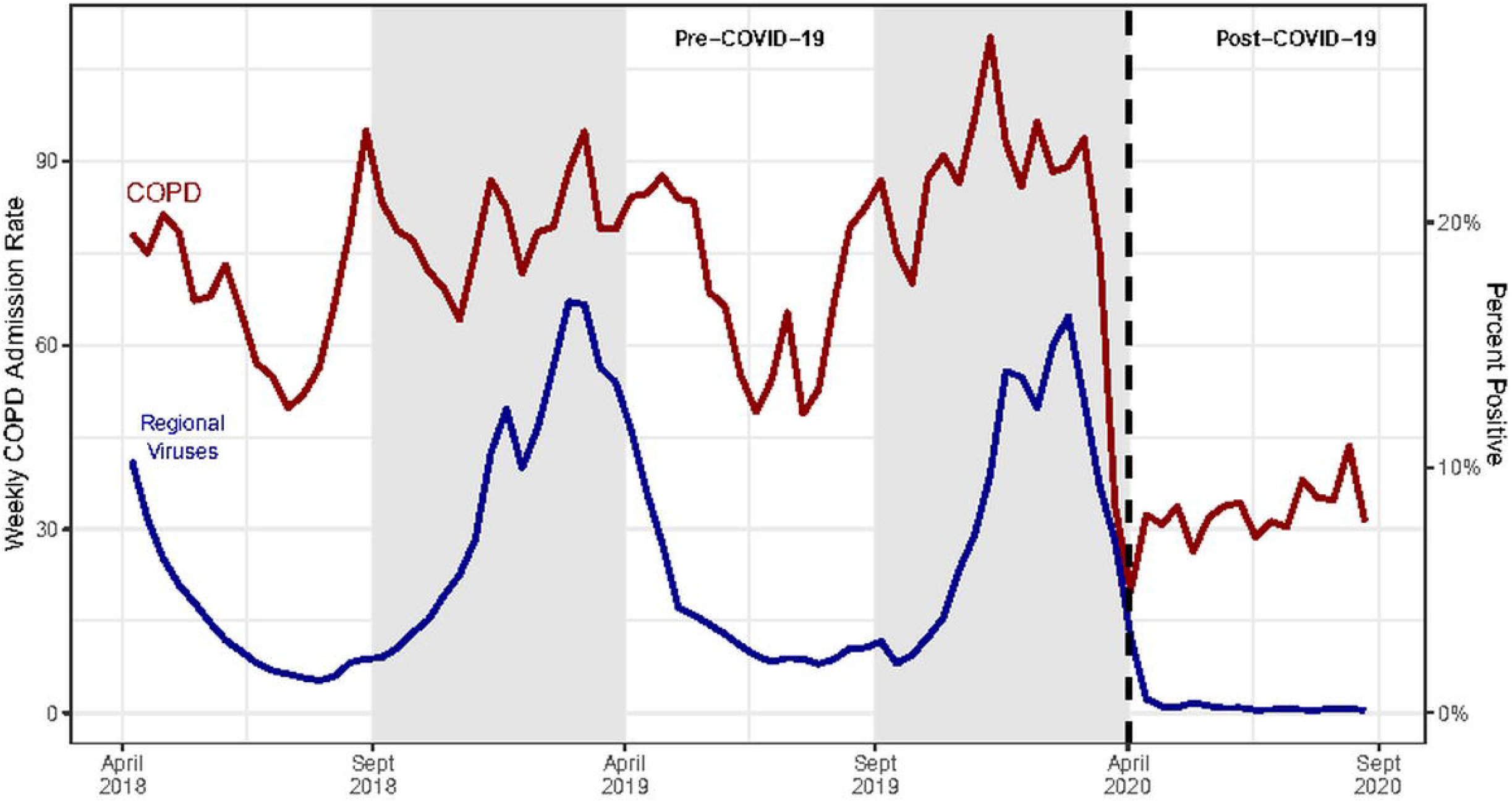
Weekly COPD Admissions Rates and the Percent Positive for Regional Non-SARS-COV2 Human Coronavirus and Statewide Human Metapneumovirus. COPD admission and viral data are reported from April 1, 2018 through September 30, 2020. Community viral testing positivity rates were obtained from the Centers for Disease Control and Prevention (CDC) for the Southern and Mid-Atlantic Regions. Viruses include influenza, parainfluenza, respiratory syncytial virus, human non-SARS coronaviruses, and human metapneumovirus. Admission rates for COPD correlated to community viral percent positivity (r=0.64, 95% CI: 0.57 to 0.69).

### Secondary Outcomes

There were no significant differences of in-hospital mortality (0.3 vs. 0.1 deaths/week pre-vs. post-COVID-19; IRR, 0.66; 95% CI, 0.20 to 2.13) or in severe COPD exacerbations requiring intensive care unit stay (3.2 vs. 2.7 exacerbations/week pre-vs. post-COVID-19; IRR 1.01; 95% CI 0.72 to 1.35) between the pre-and post-COVID-19 periods (Table 2). Similarly, there were no significant differences in median hospital length of stay for COPD admissions.

### Subgroup Analyses

The difference in COPD admissions pre- vs. post-COVID-19 period was most pronounced for patients with three or fewer comorbidities, assessed using CCI (IRR, 0.60; 95% CI, 0. 49 to 0.75) (Figure 3, Online supplement Table 5). The reduction in COPD admissions associated with the post COVID-19 period was greater for patients without recurrent admissions than for patients with recurrent admissions (IRR, 0.66; 95% CI, 0.53 to 0.81) (Online supplement Table 5).

**Figure 3.**
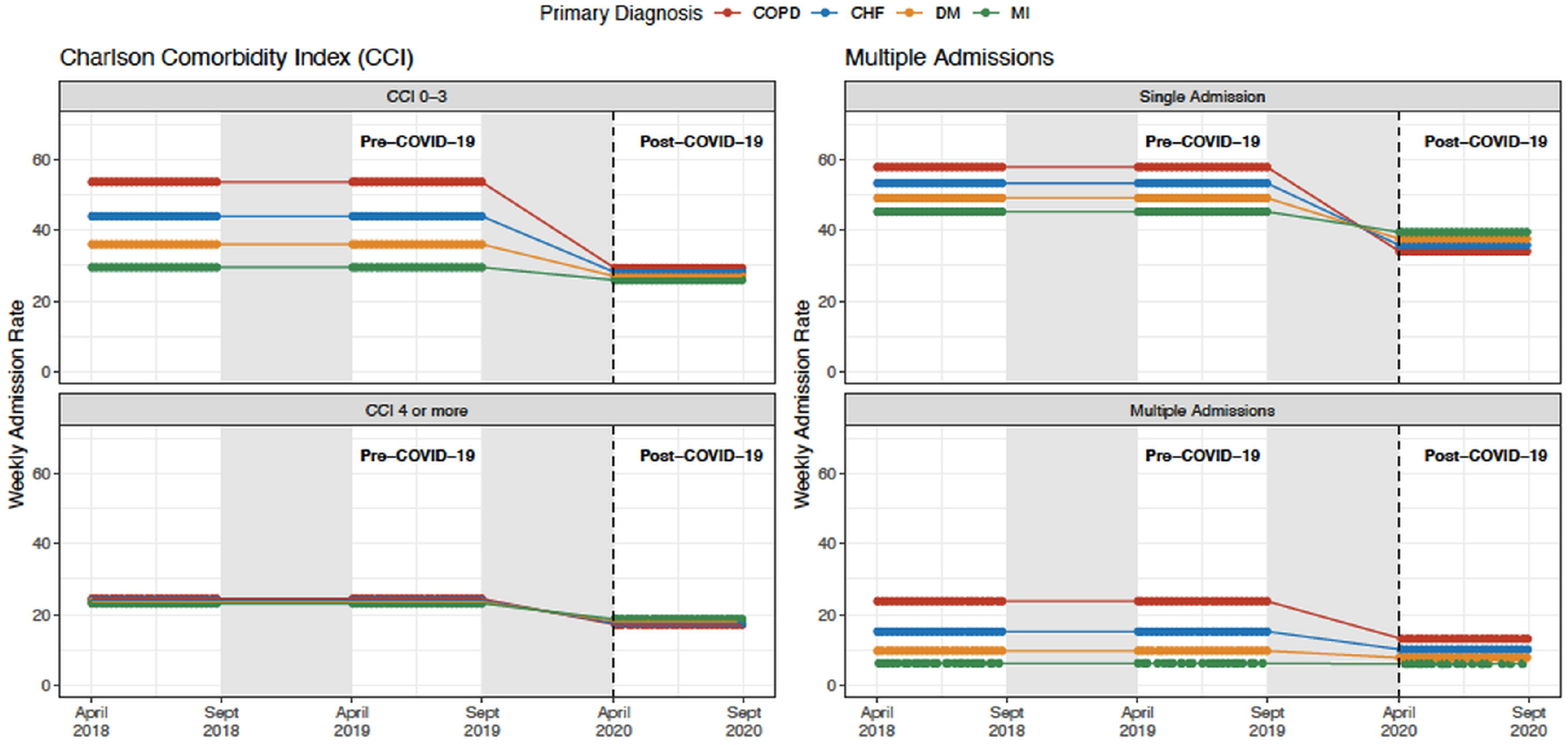
Weekly Hospital Admission Rates for Chronic Obstructive Pulmonary Disease, Congestive Heart Failure, Diabetes Mellitus, and Myocardial Infarction Stratified by Charlson Comorbidity Index and Recurrent Admissions. Our analyses compared the post-intervention period (April 1, 2020 through September 30, 2020) with a season-matched pre-intervention period comprising of April 1, 2018 through September 30, 2018 and April 1, 2019 through September 30, 2019. The grey shaded areas denote data that were not included in the season-matched analysis. Declines in COPD admission rates were most pronounced in the subgroups of patients with fewer comorbidities and without repeat admissions.

## Discussion

This study documents a substantial decline in weekly COPD admissions occurring during the COVID-19 pandemic. The reduction is greater in COPD admissions than that observed in other medical conditions less strongly associated with respiratory viral infections. This observation can be plausibly explained by reductions in community respiratory viral infections consequent to measures taken to control SARS-CoV2 transmission.

Our study includes data from a large, multihospital health system which allows us to minimize potential bias due to hospital resource allocation, such as admission limitation of non-COVID-19 patients or transfer of such patients to accommodate COVID-19 patients. All hospitals were located in a single state and, therefore, we assume more homogeneity in the observed public health measures. We included each of the 12 hospitals in the system as a random effect in our models to account for possible differences in the sociodemographics of the hospital catchments that may influence the study findings. In addition, our data included weekly admission trends dating back to 2018, allowing for a granular evaluation of changes over time. We also assessed other possible causes of the decrease in COPD admissions, including a subgroup analysis comparing patients with varying degrees of comorbidities and frequency of exacerbations, and by correlating our findings with respiratory viral trends. The difference-in-differences models was seasonally-matched and included other diagnostic conditions to account for hospital avoidance due to fear of COVID-19 transmission.

There have been prior publications that described declines in hospital admissions during the pandemic. One large study described reductions in emergency room presentations ranging from 41.5% to 63.5%, with greater reductions observed in areas more heavily affected by the pandemic.^13^ Birkmeyer et al. examined admission frequencies across a large nationwide medical group and found substantial declines to a low of 42.8% below normal. In that study, COPD admissions dropped more than any other diagnostic group to a nadir reduction of 68.6%, and despite admissions for some other conditions returning to pre-pandemic levels, COPD admissions remained 40.1% lower than baseline.^12^ A variety of mechanisms have been proposed to explain declines in medical admissions for various major non-COVID-19 conditions, but the apparently greater decline in COPD admissions has been inadequately explained.

The decrease in admissions for all causes relate to a global reduction in hospital and emergency department visits, likely due to patients’ fear of contracting SARS-CoV2 virus in these settings.^13,25,26^ There has also been a shift towards telemedicine, and more patients are treating symptoms at home than in the hospital, both of which may contribute to lowering overall hospitalization.^27,28^ However, there are additional factors that may play a role specific to the COPD population. These include improvements in air quality during the pandemic, potential increased adherence to COPD controller medications and changes in rates of smoking during the pandemic.^29 30 31^ Overall, the degree of reduction from each of these measures is likely small and fails to explain such a large, immediate reduction in COPD admissions.

As viral infections account for 40-50% of COPD exacerbations, the most likely explanation for the large decrease in COPD admissions is the reduction of respiratory virus prevalence in the population.^4,17^ Exacerbations caused by a virus also tend to be more severe and longer-lasting than non-viral exacerbations (median 13 days compared to 6 days, respectively).^17^ Recent CDC data demonstrate notable reductions of respiratory viral infections in 2020 compared to 2018 and 2019 combined.^23^ Statewide curfews, social distancing measures (including closure of indoor restaurants and limitations on large gatherings), and campaigns for frequent handwashing and mask-wearing likely contributed to this drop in respiratory viral infection prevalence.^14,15,34^ Our analysis shows strong temporal correlation between COPD admission frequencies and respiratory virus burden in Maryland (Figure 2). The decrease in respiratory virus positivity was beyond expected seasonal decrease during the post-COVID period compared to pre-COVID period (years 2018 and 2019). The correlation between respiratory virus burden and our control group conditions was substantially weaker suggesting the decrease in respiratory viral prevalence appears likely to be the greatest contributor to the decreased frequency of COPD admissions seen during the COVID-19 pandemic.

Our data also show that the reduction in COPD admissions was greatest in those with fewer comorbidities. COPD exacerbations occurring during the COVID-19 pandemic may have been associated with a patient’s underlying poor health condition at baseline. In addition, a change in engagement with the health care system during the pandemic, such as avoidance of crowded emergency department visits, may have led to a shift toward outpatient management of COPD exacerbations, especially in those with fewer comorbidities and less severe disease. Those patients with fewer comorbidities may have been able to manage exacerbations at home better than those with multiple comorbidities and baseline poor physiologic reserve. Often, these patients tend to be younger and likely explains the shift in mean age of COPD patients in the post-COVID-19 group. Older patients with more comorbidities and more severe COPD experience more frequent exacerbations, regardless of respiratory viral infections, and may not experience the decrease seen in those with fewer comorbidities.

A few limitations of our study deserve mention. Our analysis is based on hospital admission data, creating a possibility for coding variability among providers and possible misdiagnosis. However, the use of ICD codes in health services research is widely accepted, and we have employed ICD codes consistent with prior studies.^35-37^ Our data did not permit granular evaluation of whether COPD exacerbations complicated by COVID could influence ICD codes chosen, but we performed a sensitivity analysis in which the case definition of COPD exacerbation was modified to include admissions in which COVID was listed as a primary diagnosis with COPD listed as a comorbidity. Results remained significant, suggesting there was not a major issue related to misclassification bias. Another limitation involves the inclusion of control diagnoses that manifest a degree of seasonality related to community respiratory viral infections. This was an intentionally conservative approach that may underestimate the true magnitude of the reduction in COPD exacerbations attributable to respiratory viral infections. Finally, the lack of statistical significance observed in the secondary endpoints could reflect power limitations rather than a true lack of association.

## Conclusions

There is a significant decrease in weekly COPD admissions during the COVID-19 pandemic, likely due to a decrease in respiratory virus prevalence from public health measures being taken during the pandemic. There may be opportunity to judiciously apply control measures outside of pandemic conditions to reduce the burden of disease imparted by community respiratory viral infections.

## Data Availability

Data available upon request

## Author contributions

Conception and design of the study: J.Y.S., N.N.O., M.S., P.Z., R.M.R.

Analysis and interpretation of the data: J.Y.S., N.N.O, B.K., J.G.W., J.F.S., Z.Z., M.S., P.Z, C.L.B, M.D., R.M.R.

Drafting the initial draft: J.Y.S., N.N.O., R.M.R.

Reviewing and editing of the manuscript: J.Y.S., N.N.O., B.K., J.G.W., J.F.S., Z.Z., M.S, P.Z, C.L.B, M.D., R.M.R

Guarantors of the manuscript: R.M.R., J.Y.S, N.N.O

## Notes

No author reports a financial conflict of interest, real or perceived.

### Competing Interest Statement

The authors have declared no competing interest.

### Funding Statement

No funding

### Author Declarations

The study was approved by the University of Maryland institutional review board (HP-00093452).

