## Supplementary material for "Decline in COPD Admissions During the COVID-19 Pandemic Associated with Lower Burden of Community Respiratory Viral Infections": Online Supplement

**Additional Methods:**

*Rationale for control group criteria*

A difference-in-differences model estimates the average effect among an exposure group while controlling for factors unrelated to the exposure of interest that would affect both exposed and unexposed groups. For this study, that means the average association between COVID-19 public health measures on weekly COPD hospital admissions. The model requires a control group of patients that exhibit a similar pre-exposure (COVID-19 public health measures) trend. For our study, this control group comprised hospital admissions for a primary diagnosis of myocardial infarction (MI), diabetes mellitus (DM), or congestive heart failure (CHF), and were identified using ICD-10-CM codes. In addition to being common admission diagnoses in our system, these were selected for informative epidemiologic aspects. Both CHF and MI admission rates experience seasonal variability and are associated with trends in viral infections, although less robustly than observed in COPD.(1-4) As such, selection of these diagnoses was an intentionally conservative approach. Diabetes, on the other hand, demonstrates little seasonality.

*Ascertainment of Patient Characteristics*

Patient characteristics were extracted from the electronic health records to assess for any imbalance between time periods or exposure groups. We also used ICD-10-CM codes to generate Charlson Comorbidity Index scores, and diagnoses of morbid obesity, tobacco use, alcohol-related disorders, substance-related disorders, and social challenges (Online supplement, Table 1c-d).

*Additional Statistical Analysis Details*

The difference-in-differences model compared the trends in hospital admissions before (pre-COVID-19) and after public health measures related to the COVID-19 pandemic (post-COVID-19 period). The parallel trends assumption was confirmed using an equivalence test with bounds of ±2% and a negative binomial model that included an interaction between the study group and months from study onset using the pre-COVID-19 data (Online supplement, Table 2).

We performed two subgroup analyses to assess heterogeneity of effect based on co-morbidities and recurrent admissions. The comorbidities were identified at the time of admission, and reported using the Charlson Comorbidity Index (CCI) which was stratified between patients with a CCI ≤3 and patients with a CCI >3 (Online supplement 1c).(5) Patients were classified as having recurrent admissions if they had more than one admission for the same primary diagnosis during the study period. The subgroup term was interacted with the comparison groups and exposure for a difference-in-difference-in-differences model. With this approach, we accounted for the subgroup covariate as an interaction term to the difference-in-differences equation. This model ascertained if the difference-in-differences estimate changed between subgroups. Sensitivity analyses included patients with a primary admission diagnosis of COVID-19 (Online supplement 1e) to the COPD group, and our primary comparison with subsets of patients with the primary admission diagnosis of congestive heart failure, diabetes, or myocardial infarction.

Difference-in-differences estimates for the negative binomial models are reported as an incidence rate ratio with 95% confidence intervals. The difference-in-differences estimates for the quantile regression model is reported as the change in median days of hospital stay with 95% confidence intervals. Confidence intervals were calculated using bootstrap methods.

**Table 1a**: ICD-10 Codes Used for Primary COPD Admission Diagnosis.

| **COPD Category** | **ICD-10 Code** | **ICD-10-Code Description** |
| --- | --- | --- |
| Bronchitis | J40 | Bronchitis, not specified as acute or chronic |
|  | J410 | Simple chronic bronchitis |
|  | J411 | Mucopurulent chronic bronchitis |
|  | J418 | Mixed simple and mucopurulent chronic bronchitis |
|  | J42 | Unspecified chronic bronchitis |
| Emphysema | J430 | Unilateral pulmonary emphysema [MacLeods syndrome] |
|  | J431 | Panlobular emphysema |
|  | J432 | Centrilobular emphysema |
|  | J438 | Other emphysema |
|  | J439 | Emphysema, unspecified |
| COPD | J440 | Chronic obstructive pulmonary disease with (acute) lower respiratory infection |
|  | J441 | Chronic obstructive pulmonary disease w (acute) exacerbation |
|  | J449 | Chronic obstructive pulmonary disease, unspecified |

**Table 1b:** ICD-10 Codes Used for Non-COPD Primary Admission Diagnosis.

| **Comorbidity** | **ICD-10 Code** |
| --- | --- |
| Myocardial infarction | I21, I22, I252 |
| Congestive heart failure | I110, I130, I132, I255, I420, I425, I426, I427, I428, I429, I43, I50, P290 |
| Diabetes Mellitus | E080, E081, E086, E088, E089, E090, E091, E096, E098, E099, E100, E101, E106, E108, E109, E110, E111, E116, E118, E119, E130, E131, E136, E138, E139, E082, E083, E084, E085, E092, E093, E094, E095, E102, E103, E104, E105, E112, E113, E114, E115, E132, E133, E134, E135 |

**Table 1c:** ICD-10 Codes Used to Determine Charlson Comorbidity Index

| **Comorbidity** | **CCI Point** | **ICD-10 Code** |
| --- | --- | --- |
| Myocardial infarction | 1 | I21, I22, I252 |
| Congestive heart failure | 1 | I110, I130, I132, I255, I420, I425, I426, I427, I428, I429, I43, I50, P290 |
| Peripheral vascular disease | 1 | I70, I71, I731, I738, I739, I771, I790, I791, I798, K551, K558, K559, Z958, Z959 |
| Cerebrovascular disease | 1 | G45, G46, H340, H341, H342, I60, I61, I62, I63, I64, I65, I66, I67, I68 |
| Dementia | 1 | F01, F02, F03, F04, F05, F061, F068, G132, G138, G30, G310, G311, G312, G914, G94, R4181, R54 |
| Chronic pulmonary disease | 1 | J40, J41, J42, J43, J44, J45, J46, J47, J60, J61, J62, J63, J64, J65, J66, J67, J684, J701, J703 |
| Rheumatic disease | 1 | M05, M06, M315, M32, M33, M34, M351, M353, M360 |
| Peptic ulcer disease | 1 | K25, K26, K27, K28 |
| Liver disease, mild | 1 | B18, K700, K701, K702, K703, K709, K713, K714, K715, K717, K73, K74, K760, K762, K763, K764, K768, K769, Z944 |
| Diabetes without chronic complications | 1 | E080, E081, E086, E088, E089, E090, E091, E096, E098, E099, E100, E101, E106, E108, E109, E110, E111, E116, E118, E119, E130, E131, E136, E138, E139 |
| Renal disease, mild to moderate | 1 | I129, I130, I1310, N03, N05, N181, N182, N183, N184, N189, Z940 |
| Diabetes with chronic complications | 2 | E082, E083, E084, E085, E092, E093, E094, E095, E102, E103, E104, E105, E112, E113, E114, E115, E132, E133, E134, E135 |
| Hemiplegia or paraplegia | 2 | G041, G114, G800, G801, G802, G81, G82, G83 |
| Any malignancy | 2 | C00, C01, C02, C03, C04, C05, C06, C07, C08, C09, C10, C11, C12, C13, C14, C15, C16, C17, C18, C19, C20, C21, C22, C23, C24, C25, C26, C27, C28, C29, C30, C31, C32, C33, C34, C37, C38, C39, C40, C41, C43, C45, C46, C47, C48, C49, C50, C51, C52, C53, C54, C55, C56, C57, C58, C60, C61, C62, C63, C76, C801, C81, C82, C83, C84, C85, C88, C90, C91, C92, C93, C94, C95, C96, C97, C98, C99 |
| Liver disease, moderate to severe | 3 | I850, I864, K704, K711, K721, K729, K765, K766, K767 |
| Renal disease, severe | 3 | I120, I1311, I132, N185, N186, N19, N250, Z49, Z992 |
| HIV infection, no AIDS | 3 | B20 |
| Metastatic solid tumor | 6 | C77, C78, C79, C800, C802 |
| AIDS (in addition to HIV code) | 6 | B37, C53, B38, B45, A072, B25, G934, B00, B39, A073, C46, C81, C82, C83, C84, C85, C86, C87, C88, C89, C90, C91, C92, C93, C94, C95, C96, A31, A15, A16, A17, A18, A19, B59, Z8701, A812, A021, B58, R64 |

**Table 1d**: ICD-10 Codes Used to Classify Patient Characteristics.

| **Comorbidity** | **ICD-10 Code** |
| --- | --- |
| Alcohol-related disorder | F10, K70 |
| Substance-related disorder | F11, F13, F14, F15, F16, F18, F19 |
| Tobacco Use | F17 |
| Social Challenges | Z55, Z56, Z57, Z59, Z60, Z62, Z63, Z64, Z65 |
| Morbidly Obese | E6601, Z6841, Z6842, Z6843, Z6844, Z6845 |

**Table 1e**: ICD-10 Code Used to Determine Primary COVID-19 Admission Diagnosis.

| **Diagnosis Category** | **ICD-10 Code** | **ICD-10-Code Description** |
| --- | --- | --- |
| COVID-19 | U071 | COVID-19 |

**Table 2.** Test for Parallel Trends in Pre-COVID Period.

| **COPD vs.** | **Incidence rate ratio (95% CI)** | **Equivalence Test P Value** |
| --- | --- | --- |
| CHF + DM + MI | 0.996 (0.995 to 0.998) | <0.001 |
| CHF | 0.996 (0.995 to 0.997) | <0.001 |
| DM | 0.995 (0.994 to 0.996) | <0.001 |
| MI | 0.997 (0.995 to 0.998) | <0.001 |

**Table 3.** Sensitivity Analysis of COPD Admissions, Including Admissions with Primary COVID-19 Diagnosis with Secondary COPD Diagnosis.

|  | **Pre-COVID-19** | **Post-COVID-19** |  | **Difference-in-Differences** |
| --- | --- | --- | --- | --- |
| **Primary outcome: weekly admissions rates** | | |  | *Incidence rate ratio (95% CI)* |
| COPD admissions | 67.4 | 43.9 |  | 0.86 (0.79 to 0.96) |
| CHF, DM, MI admissions | 191.0 | 149.7 |  |  |

**Table 4.** Sensitivity Analysis of Weekly Hospital Admissions Using a Diagnosis-Specific Subgroups as Controls.

|  | **Pre-COVID-19** | **Post-COVID-19** |  | **Difference-in-Differences** |
| --- | --- | --- | --- | --- |
| **CHF control** |  |  |  | *Incidence rate ratio (95% CI)* |
| COPD admissions | 67.4 | 31.5 |  | 0.66 (0.59 to 0.75) |
| CHF admissions | 87.0 | 66.8 |  | Ref (1.00) |
| **DM control** |  |  |  |  |
| COPD admissions | 67.4 | 31.5 |  | 0.57 (0.51 to 0.65) |
| DM admissions | 55.3 | 47.6 |  | Ref (1.00) |
| **MI control** |  |  |  |  |
| COPD admissions | 67.4 | 31.5 |  | 0.69 (0.60 to 0.78) |
| MI admissions | 48.7 | 35.4 |  | Ref (1.00) |

**Table 5.** Subgroup Analysis of Weekly Hospital Admissions using a Differences-in-Difference-in-Differences Model.

| **Subgroup** | **COPD Admissions** | | **CHF, DM, and MI Admissions** | | **Difference-in-Differences-in-Differences** | **P- value** |
| --- | --- | --- | --- | --- | --- | --- |
|  | **Pre-COVID-19** | **Post-COVID-19** | **Pre-COVID-19** | **Post-COVID-19** |  |  |
| **Charlson Comorbidity Index** |  |  |  |  | *Incidence rate ratio (95% CI)* |  |
| 0-3 | 56.6 | 25.3 | 107.0 | 85.1 | 0.60 (0.49 to 0.75) | <0.001 |
| 4 or more | 10.8 | 6.2 | 84.2 | 64.6 | Ref (1.00) |  |
| **Recurrent admissions** |  |  |  |  | *Incidence rate ratio (95% CI)* |  |
| Only 1 admission during study period | 52.2 | 23.7 | 153.1 | 123.4 | 0.66 (0.53 to 0.81) | <0.001 |
| More than 1 admission during study period | 15.2 | 8.04 | 37.7 | 26.6 | Ref (1.00) |  |
